# *Leptospira* infection in rural areas of Urabá region, Colombia: a prospective study

**DOI:** 10.1101/2021.10.23.21265419

**Authors:** Juan C. Quintero-Vélez, Juan D. Rodas G, Carlos Rojas A, Albert I. Ko, Elsio A. Wunder

**Affiliations:** Grupo de Investigación Ciencias Veterinarias Centauro, Universidad de Antioquia, Medellín, Colombia; Grupo de Investigación Microbiología Básica y Aplicada, Universidad de Antioquia, Medellín, Colombia; Department of Epidemiology of Microbial Diseases, Yale School of Public Health, New Haven, CT, 06510; United States; Grupo de Epidemiología, Universidad de Antioquia, Medellín, Colombia; Gonçalo Moniz Institute, Oswaldo Cruz Foundation; Brazilian Ministry of Health; Salvador, BA, 40296-710; Brazil

**Keywords:** *Leptospira*, leptospirosis, Colombia, Urabá, seroprevalence, seroincidence

## Abstract

The objective of this study was to analyze the eco-epidemiological aspects of *Leptospira* seroprevalence and seroincidence, and its associated factors in two municipalities of northwest Colombia. A prospective study was performed in rural areas of Urabá, Antioquia, Colombia. The study enrolled 597 people between November-2015 and January-2016, of which 274 people were followed up one year later. Serologic testing was performed by a microscopic agglutination. The outcomes were seroprevalent and seroincident cases, and the main exposure was an outdoor occupation. A binary and mixed-effect multinomial logistic regression model was used to estimate factors associated with seroprevalent or seroincident cases of *Leptospira* infection. The overall *Leptospira* seroprevalence was 27.81% (95%CI:23.62-32.49) and the overall cumulative seroincidence for *Leptospira* was 14.60% (95%CI:10.33–20.23). Multivariable analysis showed that factors associated with *L. interrogans* serogroups seropositivity were outdoor occupation, male gender, older age, the presence of dirt soil in the household, and the presence of piglets and opossums. It also showed that factors associated with other *Leptospira* species serogroups were the presence of pit latrines and of turkeys. In addition, the multivariable model of seroincident cases of *L. interrogans* serogroups evidenced outdoor occupations, the presence of rats, and corn cultivation as risk factors. Likewise, the multivariable model for seroincident cases of other *Leptospira* species showed that the presence of hunting canines and cassava cultivation were risk factors. We found specific factors associated with the transmission of *Leptospira* serogroups contribute to the understanding of the epidemiology of Leptospira infection in rural areas of Urabá, Colombia.

**AUTHOR SUMMARY:** More than one million cases of leptospirosis occur each year, and about 60,000 people die of the disease worldwide. Leptospirosis is a zoonotic bacterial disease imparting its heaviest burden on resource-poor populations. It is also a major cause of economic loss in farm animals causing fetal infections, abortion, stillbirths, infertility, and increased culling rate. Leptospires also contaminate surface waters and survive in mud and moist soils, facilitating transmission among hosts. This complex cycle of transmission makes the disease a suitable model for a one-health approach. This is a prospective study conducted in rural areas of Colombia aimed at identifying the factors associated with seroprevalent and seroincident *Leptospira* cases. In addition, the results showed that the presence of synanthropic and domestic animals, the characteristics of household materials, and individual variables were factors associated with *Leptospira* seropositivity. This study clarifies the epidemiology of Leptospira infection and could improve the surveillance program in Colombia, especially in rural areas.

## INTRODUCTION

Leptospirosis is a zoonotic bacterial disease that burdens resource-poor populations the most ^1^. The disease is caused by pathogenic species of the genus *Leptospira*, which comprises 64 species^2^. The incidence rate of leptospirosis worldwide ranges from 0.10 to 975 cases per 100,000 population, and new cases are more frequent in tropical countries ^3,4^. Environmental (rainfall, seasons, and erosions) and ecological factors, such as the relationship between human-domestic animals and wild mammals, and socio-cultural characteristics, including outdoor occupations and practices, increase the number of leptospirosis cases ^5–8^.

Leptospirosis has been a mandatory notifiable disease in Colombia since 2007 ^9^, with a reported case fatality rate of between 1.40% and 2.24% during the period 2016–2018 ^10–12^. During the same period, 1,885 cases of the disease were confirmed; a significant proportion (18.30%) of these cases occurred in the Department of Antioquia, especially in the municipalities of Turbo and Apartadó in the Urabá region ^10–12^. In this region, the seroprevalence of *Leptospira* was estimated to be 12.50% (95% CI 10.01–15.50) in 2007, with the highest seropositivity proportion found in Carepa (27.3%), Necoclí, and San Pedro de Urabá (25.00%), followed by Apartadó (14.80%), Turbo (11.80%), and Chigorodó (7.50%) ^13^. However, higher seropositivity was found later in Necoclí (35.60%) in 2009, indicating a potential increase in the transmission in the region ^14^. Among patients with febrile syndrome studied between 2007 and 2008 in Apartadó and Turbo, 31 cases (14.1%) were diagnosed with leptospirosis, and serogroups Tarassovi and Semaranga were identified in those patients ^15^. Moreover, studies conducted in the Urabá region have identified socio-ecological factors associated with *L. interrogans* exposure ^14^.

Previous studies have shown that cases of leptospirosis are occurring in the Urabá region ^16^. The Urabá region, especially rural areas, has conditions favoring the circulation of *Leptospira* such as the presence of synanthropic rodents in households, the presence of wild animals in peridomiciliary areas, keeping domestic animals under unsanitary conditions, occupations with a potential risk of infection (agriculture), and inappropriate water supply and sewage disposal methods ^17–19^. Besides, the failure to report cases of leptospirosis in Urabá has been related to poor recognition of areas and people at risk of presenting the disease ^16,20^. However, additional analytical studies aimed identifying factors associated with the seroprevalence and seroincidence of infection with different *Leptospira* serogroups have rarely been conducted in Colombia.

The overall objective of this study was to analyze the eco-epidemiological aspects of *Leptospira* seroprevalence and the incidence and associated risk factors of infection in two municipalities of northwest Colombia. Through a prospective study analysis, we were able to identify socio-ecological factors (human, wild, and domestic animal relationships, age, occupation, land use, and household characteristics) related to *Leptospira* seropositivity in humans and determine the seroincidence of *Leptospira* infection among individuals living in those areas. Our study provides a better understanding of the epidemiology of *Leptospira* infection in the region that can be expanded to similar areas in Colombia and throughout the world. It can also help guide public health measures to help intervention strategies and reduce the transmission of leptospirosis.

## MATERIALS AND METHODS

### Study design

A prospective study was conducted in two rural areas of Urabá, Colombia, in the localities of Alto de Mulatos, Turbo (8°08’12.5”N 76°33’01.7”W), and Las Changas, Necoclí (8°32’52.5”N 76°34’23.7”W) (Figure 1) located 356 and 418 km from Medellín city, the capital city of the Department of Antioquia. Between November 2015 and January 2016, individuals of both genders residing in the study area who agreed to sign the informed consent were included in the baseline study (T0). People were excluded if they planned to move out from the study area within the following year and if they were suspected of being affiliated with illegal armed groups (this information was contributed by a research team that inhabited the study region). The same participants were followed up 12 months later, between November 2016 and January 2017 (T12).

**Figure 1.**
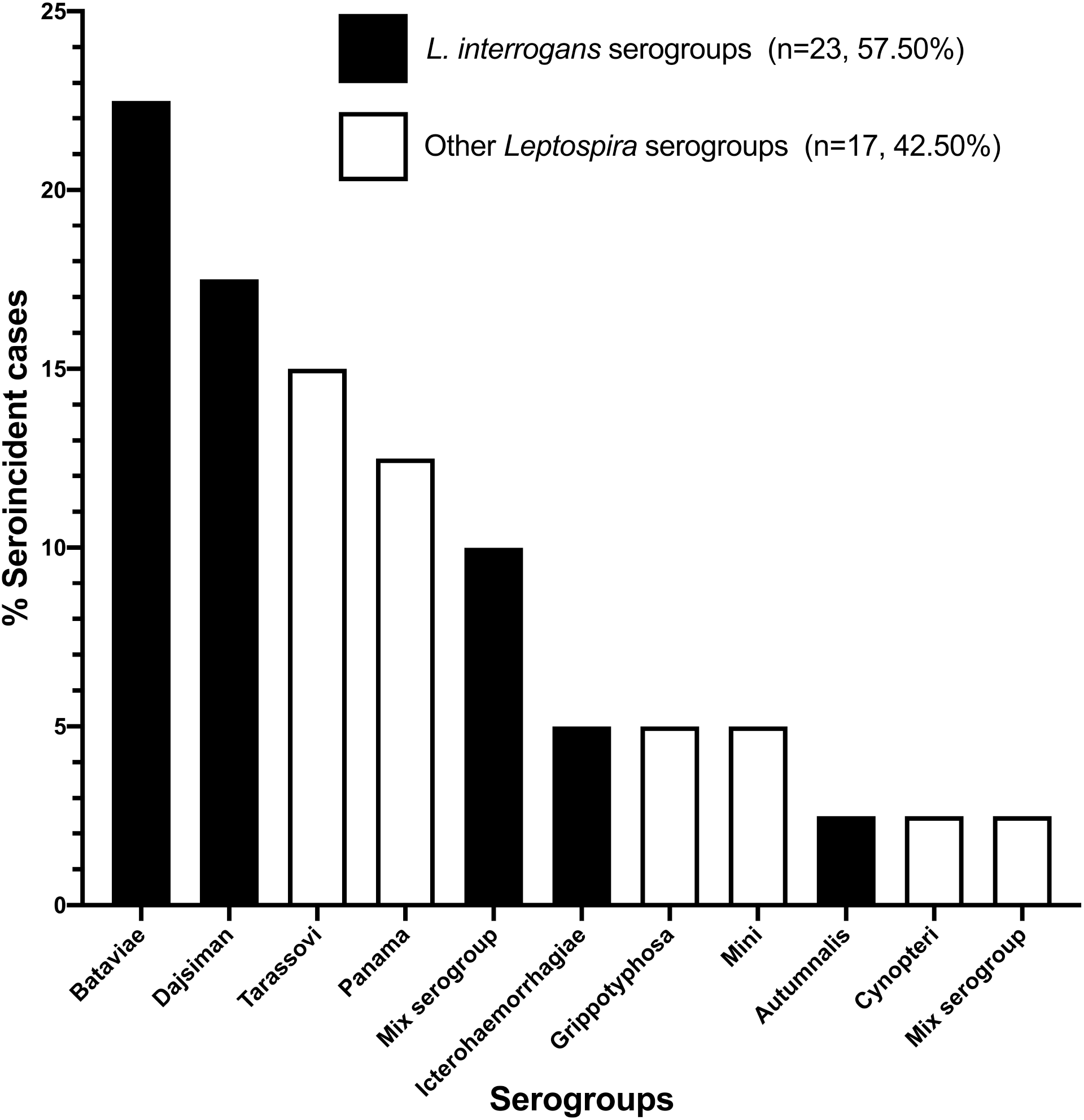
Map of the study area localized in rural areas of Urabá region of Antioquia, Colombia.

### Sampling design

Nine hamlets, five in Alto de Mulatos and four in Las Changas, were selected according to the ecological factors associated with infectious tropical diseases (the presence of wild mammals, the characteristics of the water supplies, and the presence of mosquitos and ticks, among others), the number of households, the presence of illegal armed groups, and the distance to urban centers (because of study logistics). This study constitutes a secondary prospective analysis of the main study designed to estimate the seroprevalence against agents of the *Rickettsia* genus ^21^. However, the proportion of seropositivity used for sample size estimation and the sampling design allowed other outcomes to be strongly and confidently evaluated. A complex random sampling was carried out with households as the sample units and their inhabitants as the analysis units. Based on the 461 households and 1,915 inhabitants identified in the census tract, a sample size of 208 households was estimated, using a 95% level of confidence, 5% error, and 41% expected prevalence ^21^. The sampling was proportional to the number of households in each hamlet, and a second serum sample was collected a year later from the same individuals included in the baseline study.

### *Leptospira* seropositivity

The research team collected blood samples from participants at baseline (T0) and the followed-up study (T12). The serum samples were tested by microscopic agglutination test (MAT). To select the antigens for sample testing, a panel of 32 *Leptospira* strains, representing different species and serogroups, was evaluated (Table 1) using a random selection of 20% of the total human serum samples collected at T0. After the evaluation, antigens with no agglutination results were eliminated from the panel, and a final list of 16 antigens was established to be tested with all sera for this study (Table 1). The MAT assay was performed according to Felzemburgh, *et al*. ^6^. The evaluation and final screening assays were performed using dilutions of 1:50 and 1:100 for human samples, and a positive sample was determined when 50% or more leptospires for a specific strain were agglutinated. Following the screening assay, positive samples were titrated to determine the higher titer for each strain.

**Table 1.** Panel of *Leptospira* strains used in this study for the microagglutination test (MAT)

| Species | Serogroup | Serovar* | Strain |
| --- | --- | --- | --- |
| *L. interrogans* | **Djasiman** | **Djasiman\*** | **Djasiman** |
| *L. interrogans* | Icterohaemorrhagiae | Icterohaemorrhagiae | RGA |
| *L. interrogans* | Icterohaemorrhagiae | Copenhageni | M 20 |
| *L. interrogans* | **Icterohaemorrhagiae** | **Copenhageni\*** | **L1 130** |
| *L. interrogans* | Sejroe | Hardjo | Hardjoprajitno |
| *L. interrogans* | Sejroe | Wolffi | 3705 |
| *L. interrogans* | **Bataviae** | **Bataviae\*** | **Van Tienen** |
| *L. interrogans* | **Pyrogenes** | **Pyrogenes\*** | **Salinem** |
| *L. interrogans* | **Pyrogenes** | **Manilae\*** | **L495** |
| *L. interrogans* | **Pomona** | **Pomona\*** | **Pomona** |
| *L. interrogans* | **Autumnalis** | **Autumnalis\*** | **Akiyami A** |
| *L. interrogans* | **Canicola** | **Canicola\*** | **H. Utrecht IV** |
| *L. interrogans* | Hebdomadis | Hebdomadis | Hebdomadis |
| *L. interrogans* | **Australis** | **Bratislava\*** | **Jez Bratislava** |
| *L. noguchii* | Louisiana | Louisiana | LSU 1945 |
| *L. noguchii* | **Panama** | **Panama\*** | **CZ 214 K** |
| *L. borgpetersenii* | **Tarassovi** | **Tarassovi\*** | **Perepelitsin** |
| *L. borgpetersenii* | Ballum | Castellonis | Castellon 3 |
| *L. borgpetersenii* | Ballum | Ballum | Mus 127 |
| *L. borgpetersenii* | **Mini** | **Mini\*** | **Sari** |
| *L. weilii* | Celledoni | Celledoni | Celledoni |
| *L. weilii* | Javanica | Coxi | Cox |
| *L. kirschneri* | **Cynopteri** | **Cynopteri\*** | **3522C** |
| *L. kirschneri* | **Grippotyphosa** | **Grippotyphosa\*** | **Duyster** |
| *L. santarosai* | Shermani | Shermani | 1342 K |
| *L. santarosai* | **Tarassovi** | **ND\*** | **AIM** |
| *L. santarosai* | Tarassovi | ND | JET |
| *L. alexanderi* | Manhao | Manhao 3 | L 60T |
| *L. alstoni* | Ranarum | Pingchang | 80-412T |
| *L. kmetyi* | **Tarassovi** | **Malaysia\*** | **Bejo-Iso9** |
| *L. mayottensis* | ND | ND | 200901122 |
| *L. biflexa* | Semaranga | Patoc | Patoc 1 |

\*Final selection of strains used for MAT evaluation of all human serum samples

### Outcome definitions

In the primary analysis, the outcome of seroprevalent cases of *Leptospira* infection was defined as a positive MAT titer ≥1:50 with 50% or more leptospires agglutination at baseline (T0). Also, the outcome of seroincident cases of *Leptospira* infection was measured by MAT seroconversion from negative to positive or by a minimum four-fold increase in titers between follow-up samples (T12 vs. T0). The serogroup that represented the highest antibody titer was defined as the presumptive infecting serogroup. However, when two or more serogroups had the same titer, the sample was considered mixed serogroup seropositivity. Additionally, the seroprevalent and seroincident outcomes were categorized as seropositive to *Leptospira interrogans* serogroups and seropositive to other *Leptospira* species serogroups to perform a secondary analysis (multinomial analysis) (Table 1).

### Study exposures and covariates

The main exposure was by working outdoors (farmers, ranchers, agricultural workers, day laborers, and water recollectors, among others). For the T0 study, this exposure was analyzed as a previous outdoor occupation (within the last five years) and a recent outdoor occupation (the previous year before the sample collection). For the prospective study, the outdoor occupation was evaluated based on the previous year before the sample collection.

Additionally, individual variables such as age (years), gender, ethnicity, time of residence in the study area, educational level, and previous episodes of fever (any past episode and during the last year of follow-up) were evaluated as potential confounders of the association between the main exposure and the outcomes.

Secondary exposures were access to public services (the presence of indoor plumbing, waste disposal, the presence of sewage, the presence of latrines, and the presence of pit latrines), the presence of domestic animals in intra or peridomiciliary areas (the presence of canines, felines, poultry, turkeys, pigs, horses, donkeys, and mules), and the presence of synanthropic or wild animals in peridomiciliary areas (the presence of rodents and opossums). Additionally, covariates such as the type of roof and floor, wall materials, the characteristics of peridomiciliary areas (the presence of bushes, trees, grasses, corn cultivation, cassava cultivation, and tomato cultivation), household location (urban center and rural area) and household proximity (very near, near, scattered, and very scattered) were analyzed. Finally, common practices among family members such as forest fragmentation, deforestation, and the use of any rodent elimination measures were considered as covariates.

### Statistical analysis

The seroprevalence and cumulative seroincidence were estimated considering the number of seropositive cases in the numerator and the number of people evaluated in both times (T0 and T12) as a denominator. Also, the confidence intervals were adjusted by random effects of the models (hamlets) for seroprevalence and seroincidence. In addition, the seroprevalent and seroincident cases were characterized using relative and absolute frequencies for qualitative variables and median and interquartile range for quantitative variables.

To estimate risk factors for seroprevalent and seroincident cases of *Leptospira* infection, a mixed-effects binary logistic regression model was used (seronegative vs seropositive samples). Additionally, a multinomial analysis was conducted using a mixed-effects multinomial logistic regression model for a specific evaluation of factors associated with the outcome of seropositive against *L. interrogans* serogroups and seropositive against other *Leptospira* species serogroups. Moreover, both models included three levels: individuals within households, households within hamlets, and hamlets (a random effect model using a variance component correlation matrix). The association between the main outcome (*Leptospira* seropositivity in humans) and variables at each level was evaluated (household and hamlet level). The linearity assumption was confirmed previous to the inclusion of quantitative variables in bivariate and multivariate models. Variables included in the multivariate models were those with p < 0.25 in bivariate analysis. The multivariate analyses were performed using the stepwise method based on the researcher’s criteria. The multilevel models of risk factors for *Leptospira* seropositivity in humans were weighted by the inverse probability of human selection in each hamlet.

Confounders were evaluated in multivariate models, and the model that best explained the outcome was selected according to Bayesian Information Criteria (BIC). Odds ratios (OR) were estimated in the cross-sectional study, and relative risks (RR) were estimated in the prospective study according to Localio, 2007 ^22^. All analyses were performed in SAS 9.04.01 using PROC GLIMMIX [17].

### Ethics statement

The Committee of Ethics in Research of the University of Antioquia approved all procedures carried out in the present study, and participants were enrolled according to written informed consent procedures previously approved by the Committee of Ethics in Research.

## RESULTS

### *Leptospira* seroprevalence and associated factors

#### Binary outcome analysis

For the baseline study (T0), 597 individuals (255 from Alto de Mulatos and 342 from Las Changas) that inhabited 246 households located in the rural areas (18% of oversampling) were enrolled. One hundred and three of them were in Alto de Mulatos and 143 in Las Changas. The sampling coverage of households and persons were 100% and 58.41% (597/1022), respectively. The overall *Leptospira* seroprevalence was 27.81% (166/597) (95% CI: 23.62-32.49), and the socio-ecological characteristics showed that 42.77% (71/166) of seropositive individuals had outdoor occupations, 52.41% (87/166) identified themselves as of male gender, and the mean age in years was 31.43 (IQR: 18.94-48.72). Additionally, among the seropositive individuals, 62.65% (104/166) reported a history of fever, and 79.52% (132/166) reported inhabiting houses with dirt soil floors. Regarding the presence of domestic animals in intra or peridomiciliary areas, 55.42% (92/166) of seropositive individuals reported having seen opossums in the peridomiciliary, area and 26.51% (44/166) reported the presence of piglets in their household (Table S1).

Our analysis identified outdoor occupations (OR = 1.56; 95% CI:1.05–2.31), the presence of a dirt soil floor in the household (OR = 1.62; 95% CI:1.14-2.31), the presence of opossums in the peridomiciliary area (OR = 1.34; 95% CI:1.003–1.80), and the tenancy of piglets (OR = 1.45; 95% CI:1.02-2.07) as risk factors to seropositivity against *Leptospira*. Also, the age as a categorized variable (OR _> 15–29 years vs ≤ 1-15 years_ = 2.30; 95% CI:1.47–3.60 and OR _> 46 years vs ≤ 1–15 years_ = 1.95; 95% CI:1.07-3.55), the male gender (OR = 2.01; 95% CI:1.42–2.84) and a history of fever (OR = 1.42; 95% CI:1.05-1.91) were factors associated with seropositivity (Table S1).

#### Multinomial outcome analysis

According to our multinomial analysis, the seroprevalence of *L. interrogans* serogroups was 18.25% (109/597) (95% CI: 14.34-23.11), and the seroprevalence of other *Leptospira* serogroups was 9.56% (57/597) (95% CI: 7.11–12.70). Among the participants seropositive to *L. interrogans*, the most frequent serogroup was Bataviae, followed by mixed serogroups and Djasiman. In addition, among the participants seropositive to other *Leptospira* species, the most frequent serogroup was Tarassovi, followed by Panama and Cynopteri (Figure 2).

**Figure 2.**
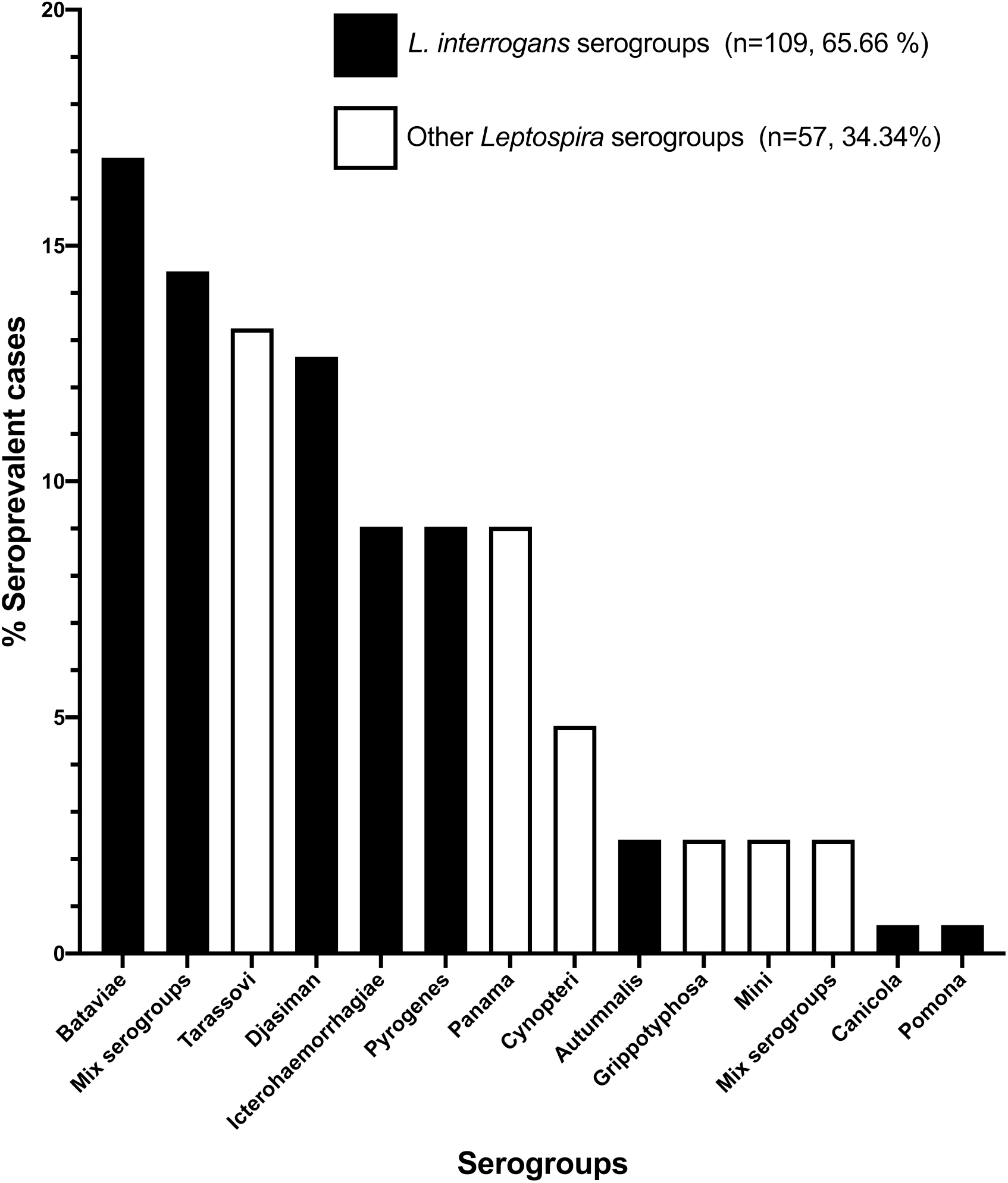
Frequency of *L. interrogans* and other *Leptospira* serogroups in the seroprevalent cases.

Sociodemographic characteristics showed that 58.72% (64/109) of seropositive individuals against *L. interrogans* and 40.35% (23/57) seropositive for other *Leptospira* species were male. The median age of seropositive individuals was 33.23 years (IQR:19.67-49.25) and 28.42 years (IQR: 16.76–44.66) among *L. interrogans* and other *Leptospira* species, respectively. Of note, a higher proportion of the seropositive to *L. interrogans* serogroups had an outdoor occupation (52.29%, 57/109), while only 24.56% (14/57) of seropositive persons to other *Leptospira* species serogroups had similar occupations (Table S2).

Most of the participants that were seropositive to *L. interrogans* (81.65%, 89/109) and other *Leptospira* species (75.44%, 43/57) inhabited households with dirt soil floors. Approximately half of the seropositive individuals in both groups had indoor plumbing (49.54% for *L. interrogans* and 54.39% for other *Leptospira* species). Additionally, the presence of pit latrines in the households was 45.78% (52/109) for people with *L. interrogans* seropositivity and 47.37% (27/57) for other *Leptospira* species seropositivity (Table S2).

Regarding the presence of domestic, synanthropic, and wild animals, 60.55% (66/109) of people with seropositivity against *L. interrogans* and 45.61% (26/57) of people with seropositivity against other *Leptospira* species reported the presence of opossums in the peridomiciliary area. The presence of rats in intra and peridomiciliary areas was 84.40% (92/109) and 85.96% (49/57) in people seropositive against *L. interrogans* and other *Leptospira* species, respectively. In addition, 1.83% (2/109) of people seropositive to *L. interrogans* and 7.02% (4/57) of people seropositive to other *Leptospira* species had hunting canines (Table S2).

The multivariable multinomial analysis showed that occupations such as ranchers, daily laborers, farmers, and agricultural (and similar) workers were a risk factor for the outcome of *L. interrogans* serogroups seropositivity (OR = 2.06; 95% CI:1.31-3.26). In addition, the male gender (OR = 2.34; 95% CI:1.55–3.53) and older age (OR _> 15–29 years vs ≤ 1-15years_ = 2.52; 95% CI:1.44–4.41 and OR _> 46 years vs ≤ 1-15yeras_ = 1.95; 95% CI:1.07–3.55), the presence of dirt soil in the household (OR = 1.87; 95% CI:1.21-2.89), and the presence of piglets (OR = 1.66; 95% CI:1.11–2.49) and opossums (OR = 1.58; 95% CI:1.11-2.23) in the peridomiciliary area were factors associated with *L. interrogans* serogroups seropositivity. Besides, the multivariable model for the outcome of seropositivity to other *Leptospira* species serogroups adjusted by occupation, gender, and age showed that the presence of pit latrines (OR = 2.34; 95% CI:1.27– 4.32) and turkeys as companionship (OR = 4.99; 95% CI:2.03-12.30) were risk factors (Table 2).

**Table 2.**
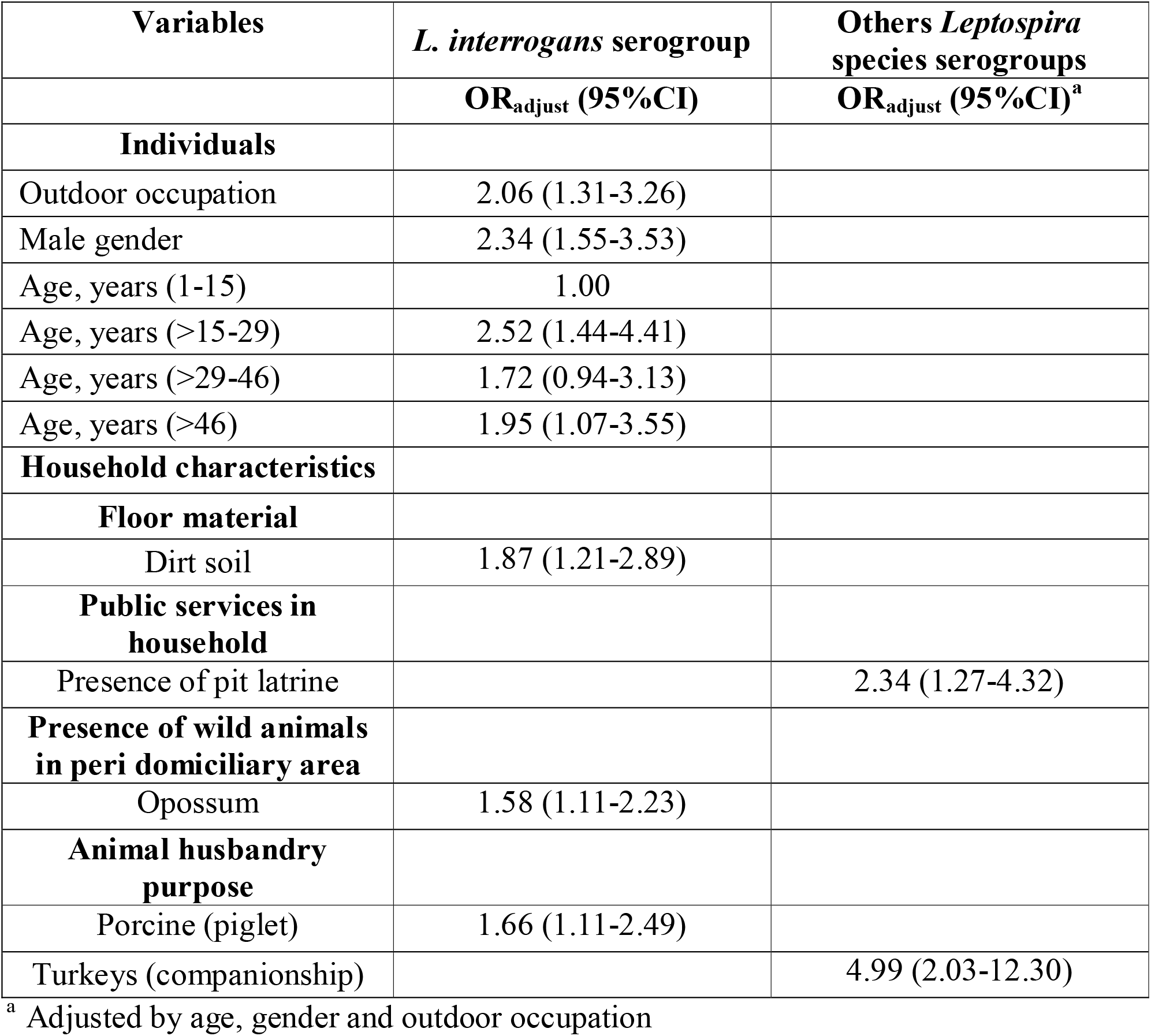
Multivariate analysis of seroprevalent cases of *Leptospira* among different serogroups (mixed-effects multinomial logistic regression model)

### *Leptospira* seroincidence and associated factors

#### Binary outcome analysis

In the follow-up study, 274 participants were enrolled at 12 months (T12), and 120 and 154 people were followed up in Alto de Mulatos and Las Changas, respectively. In Table S3, the characteristics of the people included in the study and lost to the follow-up in both areas are shown. The overall cumulative seroincidence of *Leptospira* was 14.60% (40/274) (95% CI: 10.33–20.23), 35.00% (14/40) of the seroincident cases had an outdoor occupation, and 27.50% (11/40) were identified as male. In addition, the median age in years of seroincident cases was 40.30 (IQR: 20.46-55.34), and 80.00% (32/40) were inhabitants of a household with a dirt soil floor. Furthermore, among the seroincident cases, 5.00% (2/40), 2.99% (3/40), and 85.00% (34/49) of the seroincident cases disclosed the presence of corn cultivation around the household and hunting canines and rats in the intra or peridomiciliary areas, respectively (Table S4). According to risk factors associated with seroincident cases of *Leptospira* infection, an outdoor occupation in the last year (RR = 2.66; 95% CI:1.46–4.68), a dirt soil floor in the household (RR = 1.77; 95% CI:1.01-3.06), the presence of corn cultivation in the peridomiciliary area (RR = 8.33.74; 95% CI:2.21–19.65), the presence of hunting canines (RR = 5.16; 95% CI:1.83-10.74), and rat infestation (RR = 2.00; 95% CI:1.03–3.76) in the intra or peridomiciliary area were identified (Table S4).

#### Multinomial outcome analysis

The multinomial analysis regarding serogroups showed that the seroincidence for *L. interrogans* serogroups and other *Leptospira* species serogroups was 8.39% (23/274) (95% CI: 5.25-13.15) and 6.20% (17/274) (95% CI: 3.38-10.93), respectively. Seroincident cases of *L. interrogans* and other *Leptospira* serogroups were male in 30.43% (7/23) and 23.53% (4/17) of cases, respectively. Also, 39.13% (9/23) of seroincident cases of *L. interrogans* serogroups and 29.41% (5/17) of seroincident cases of other *Leptospira* serogroups had had outdoor occupations in the previous year. The median age for the seroincident cases was 47.46 years (IQR: 24.76-58.42) and 29.99 years (18.19–49.28) for *L. interrogans* serogroups and other *Leptospira* species serogroups, respectively. Only one person in the group of *L. interrogans* reported a history of fever in the year of follow-up (Table S5).

In addition, for the characteristics of the peridomiciliary area, 8.70% (2/23) of seroincident cases by *L. interrogans* serogroups reported corn cultivations around their houses. Cassava cultivations were also reported in the peridomiciliary area of seroincident cases for *L. interrogans* (13.04%, 3/23) and other *Leptospira* serogroups (17.65%, 3/17). Additionally, households of seroincident cases for both outcomes had a high frequency of rat infestation with 91.30 % (21/23) for *L. interrogans* and 76.47% (13/17) for other *Leptospira* species. Only the seroincident cases for other *Leptospira* species serogroups reported hunting canines (17.65%, 3/17) (Table S5).

The multivariable model evidenced outdoor occupations in the last year (RR = 2.93; 95% CI:1.36-6.16), the presence of rats (RR = 2.81; 95% CI:1.07-7.12), and corn cultivation around the households (RR = 22.74; 95% CI:5.75-51.74) as risk factors for the seroincident cases of *L. interrogans* serogroups. Besides, the age in years (as a quantitative variable) was a risk marker (RR = 1.02; 95% CI:1.01–1.04), and male gender was a protector marker (RR = 0.34; 95% CI:0.15-0.80) for the seroincident cases of *L. interrogans* serogroups (Table 3). Likewise, the multivariable model for seroincident cases of other *Leptospira* species adjusted by occupation, age, and gender, showed that the presence of hunting canines (RR = 6.17; 95% CI:2.72–10.99) and cassava cultivation (RR = 3.05; 95% CI:1.24-6.52) were risk factors (Table 3).

**Table 3.** Multivariate analysis of seroincident cases of *Leptospira* among different serogroups (mixed-effects multinomial logistic regression model)

| **Variables** | ***L. interrogans* serogroup** | **Others *Leptospira* species serogroups** |
| --- | --- | --- |
|  | **RR<sub>adjust</sub> (95%CI)<sup>a</sup>** | **RR<sub>adjust</sub> (95%CI)<sup>b</sup>** |
| **Individuals** |  |  |
| Outdoor occupation | 2.93 (1.36-6.16) |  |
| Age (years) | 1.02 (1.01-1.04) |  |
| **Peri domiciliary area characteristics** |  |  |
| **Vegetation** |  |  |
| Corn culture | 22.74 (5.75-51.74) |  |
| Cassava culture |  | 3.05 (1.24-6.52) |
| **Synanthropic mammals** |  |  |
| Rats | 2.81 (1.07-7.12) |  |
| **Animal husbandry purpose** |  |  |
| Hunting canines |  | 6.17 (2.72-10.99) |

<sup>a</sup> Adjusted by gender
<sup>b</sup> Adjusted by age, gender and outdoor occupation

Among *L. interrogans* seroincident cases, the most frequent serogroup seropositivity was against Bataviae, followed by Djasiman and mix serogroups (Figure 3). Of note, 100% of seroincident cases of Djasiman, Icterohaemorrhagiae, and Autumnalis serogroups were exposed to rodents in their households, and 88.88% of seroincident cases to Bataviae serogroup had the same exposure to rodents. Among seroincident cases to other *Leptospira* species serogroup, the most frequent seropositivity was against Tarassovi, followed by Panama (Figure 3). Furthermore, 50% (3/6) and 60% (3/5) of seroincident cases to Tarassovi and Panama serogroups had pigs and opossums around their households, respectively.

**Figure 3.**
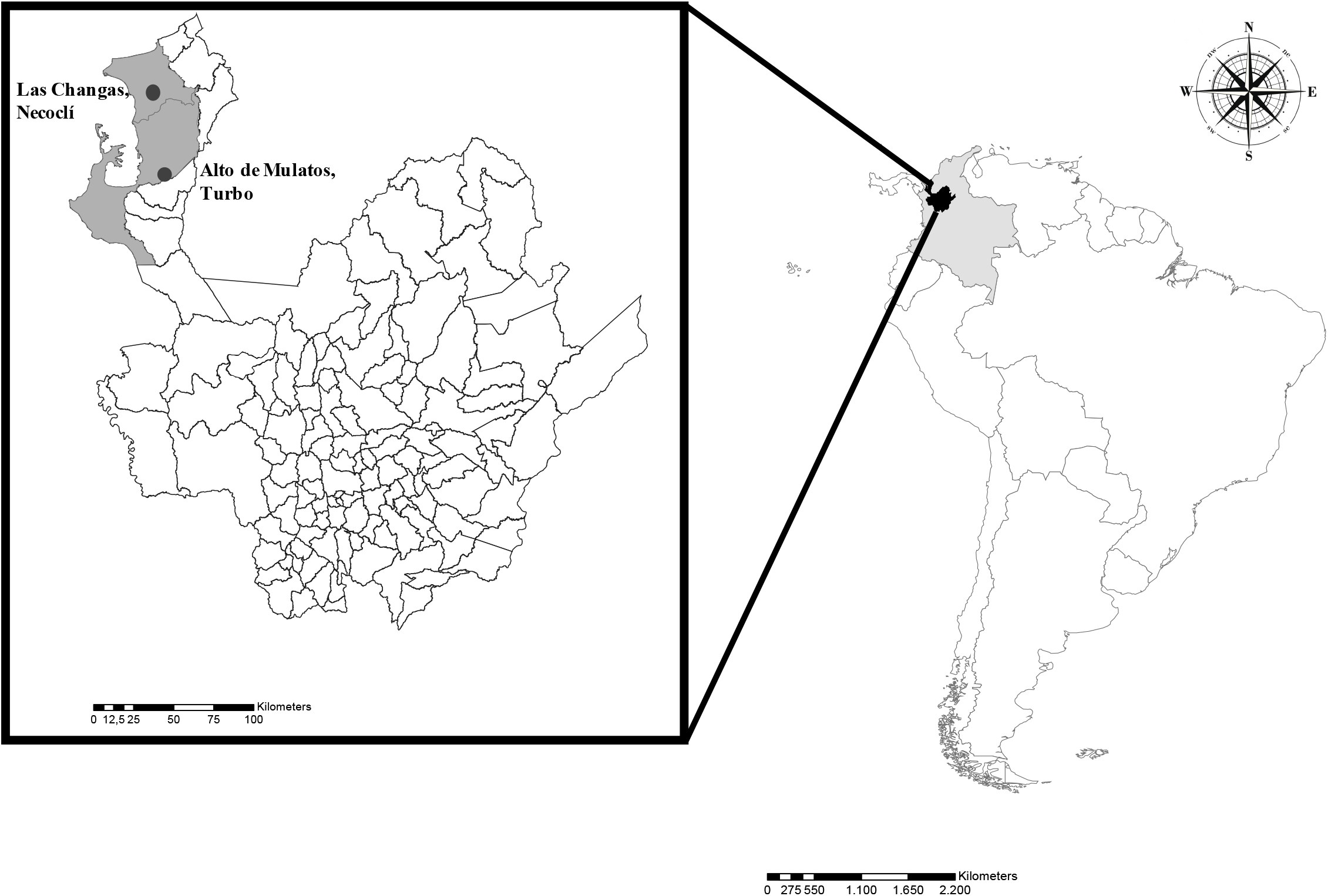
Frequency of *L. interrogans* and other *Leptospira* serogroups in the seroincident cases.

## DISCUSSION

Leptospirosis is a zoonotic disease with mandatory notification to the National Surveillance System in Public Health in Colombia since 2007. However, the program fails to detect active cases of leptospirosis in rural areas due to the low access to diagnostic tests and the lack of medical presumption ^20^. Thus, conducting prospective studies to analyze and detect potential areas at risk of transmission of tropical diseases, as well as estimating risk and protective factors for subsequent interventions, are becoming increasingly necessary. The Urabá region in the Department of Antioquia, Colombia, is considered a potential endemic region for tropical diseases such as malaria, leishmaniasis, dengue viruses, leptospirosis, and rickettsiosis, among others ^15,23,24^. Therefore, analyzing seropositivity outcomes for more than one disease when conducting analytical observational studies could broaden the understanding of factors for those tropical and neglected diseases potentially endemic in this region.

The overall *Leptospira* seroprevalence estimated in the current study was 27.81%, very similar to the report from San Juan, Puerto Rico (27.2%, 55/202) ^25^. Additional studies conducted in the Urabá region using a definition of seropositive outcome ≥ 1:100 titers by MAT, reported a *Leptospira* seroprevalence of 35.6% ^14^ and 12.5% ^13^. We also estimated an overall cumulative seroincidence of *Leptospira* of 14.60%; to the best of our knowledge, this could be the first estimation of *Leptospira* seroincidence in Colombia, which helps to identify potential areas at risk of leptospires transmission.

The factors associated with both seroprevalent and seroincident cases of *Leptospira* infection, after controlling for potentially confounding variables and adjusting for our design effect, were history of outdoor occupation such as farming, agriculture, or daily laboring, and older age. For the seroprevalence analysis, male gender was also a factor associated with *Leptospira* infection. These exposures (outdoor labor) are recognized as an occupational risk for *Leptospira* infection ^3,26^. These results are confirmed by our demographic data, showing that male individuals over 15 years old have a high probability of working outdoors in the Urabá region. Furthermore, the risk markers identified in our study, such as gender and age, were also reported as factors associated with *Leptospira* infection in previous studies ^6,27^.

Other characteristics found in our study area, such as poverty and some cultural practices (e.g., an abundance of domestic animals and precariousness in household construction) are conditions highly associated with the presence of a large number of tropical zoonotic diseases ^28^. Our analysis showed that the tenancy of piglets and the eyesight of opossums in the peridomiciliary area were important risk factors for seroprevalent cases, and the presence of a dirt floor inside households was associated with both seroprevalent and seroincident cases of *Leptospira* infection. Other studies have identified the presence of piglets and opossums in the peridomiciliary area as risk factors for *Leptospira* infection; these mammals have been considered as potential hosts participating in the life cycles of several *Leptospira* species ^3,29,30^. Also, Bierke *et al*. ^31^ found that *Leptospira* can survive for long periodsin several types of soil depending on its physical, chemical, and biological conditions such as pH, temperature, or mineral concentration. Furthermore, a significant amount of *Leptospira* has been detected in 31% (22/70) of samples from different soil microenvironments in a neighborhood endemic leptospirosis in Salvador, Bahia, Brazil ^32^.

*Leptospira* infection in the Department of Antioquia is most commonly caused by serogroups of the *L. interrogans* species ^9^. A study conducted in the Urabá region on patients with leptospirosis showed a high proportion of infection for *L. interrogans* serogroups of Copenhageni (28.6%), Hardjo (20.6%), Australis (17.5%), Canicola (11.1%), and Pomona (6.3%). Other Leptospira serogroups were also found to have a high infection proportion, including Tarassovi (25.4%), Grippotyphosa (17.5%), Shermani (9.5%), Ballum (7.9%), Cynopteri (3.2%), and Panama (1.6%) ^28^. Moreover, in the same study, of 199 patients with febrile syndrome, 11.56% were infected by *L. interrogans* serogroup Icterohaemorrhagiae, 10.05% were infected by Sejroe, and other serogroups registered a proportion of infection near to 10% ^20^. Regarding seroprevalent cases in the Urabá region, it was reported that 87% of seroprevalent cases had high titers against Icterohaemorrhagiae, followed by the Sejroe serogroup [14]. Likewise, in a study conducted in the city of Cali in the south of Colombia, the seroprevalence of *Leptospira* was 12.53%, and the most frequent serogroup in seroprevalent cases was Australis (61.4% of prevalent cases) [33].

Although multinomial regression modeling is infrequently used to analyze infectious diseases ^33^, this method had the advantage of estimating the simultaneous effects of multiple factors. In our study, we analyzed the factors associated with the risk of *Leptospira* infection, stratified in two categories (*L. interrogans* serogroups and other species serogroups), and, according to our knowledge, this is the first publication that reports its use in the study of *Leptospira* infection. In this sense, outdoor occupations, age in years, male gender, the presence of dirt soil floors, and the presence of piglets and opossums in peridomiciliary areas were factors associated with seroprevalent cases of *L. interrogans* serogroups. Additionally, the presence of pit latrines around households and turkeys as companionship were risk factors for seropositivity against other *Leptospira* species serogroups. Similarly, a lack of toilets inside the households was a factor associated with *Leptospira* seropositivity (OR unadjusted = 3.75; 95 CI%: 1.33-10.60) in a study carried out in the south of Colombia ^34^. Recently, a study in Germany identified the presence of poultry in peridomiciliary areas as a risk factor for *Leptospira* infection ^26^. However, it is most likely that the presence of poultry or turkeys in peridomiciliary areas is associated with the presence of synanthropic rodents or wild mammals, a necessary condition to maintain leptospires in the environment ^35^.

Regarding seroincidence cases, the multinomial analysis identified the presence of rodents in intra or peridomiciliary areas, outdoor occupations, the presence of corn cultivation in peridomiciliary areas (proxy of the presence of synanthropic and wild mammals), and age in years as factors associated with seroincident cases of *L. interrogans* serogroups. In this multinomial analysis, we also found that the presence of hunting canines and cassava cultivation in peridomiciliary areas were risk factors for seroincident cases for other *Leptospira* species serogroups. Similar to urban areas of Brazil, where only *L. interrogans* serogroup is circulating, household conditions (household income), sociodemographic aspects (age, gender, and occupation), and rat infestation were risk factors for *Leptospira* infection ^5,6,27^. Regarding the canines, these domestic animals have been considered important reservoirs or hosts in the infection cycle of *Leptospira* because they can expel the leptospires through their urine for long periods of time (weeks or months) ^36^. Different serogroups of *Leptospira* such as Canicola, Autumnalis, Australis, Grippotyphosa, Harjo, Pomona, Pyrogenes, and Djasiman, among others ^37–40^, have been found in canines. In Brazil, a longitudinal study of dogs found a seroprevalence between 9.3% (95% CI:6.7-12.6) and 19% (95% CI:14.1–25.2) in different periods. Additionally, an overall cumulative seroincidence of 11% (95%CI:9.1-13.2) was estimated, and the cumulative seroincidence by trimester varied between 6% (95% CI:3.3–10.6) and 15.3% (95% CI:10.8-21.2) ^40^.

In this study, reactivity to Bataviae was the most frequent among prevalent and incident cases of *L. interrogans* seropositive individuals, indicating that these cases had probably been exposed to infected rodents or canines ^41,42^. Nevertheless, detection of antibodies against Djasiman and Icterohaemorrhagiae serogroups evidenced additional exposure to sources contaminated with leptospires shed by synanthropic or wild mammals ^3,29^. Regarding different species of *L. interrogans* serogroups, this study showed a high frequency of seroprevalent and seroincident cases against the Tarassovi serogroup, which is mainly found in pigs ^3,30^. Even though the Tarassovi serogroup is present in different species of *Leptospira* considered as pathogens and classified into subgroup II, this serogroup has not been incriminated in clinical cases of leptospirosis in humans yet ^43^. The Panama serogroup was another important serogroup found in seroprevalent and seroincident cases. This serogroup was initially described in opossums, wild rodents, and numerous domestic animals. However, there are several clinical cases of leptospirosis described in humans, potentially caused by the Panama serogroup ^44,45^. Finally, reactivity to the Cynopteri serogroup was the third in seroprevalent cases and the fourth in seroincident cases. This serogroup is frequent in bats and various wild animals ^3,46,47^.

The main limitation of this study was the moderate sampling coverage of individuals (58.41%). Some participants declined the study enrollment, and others moved out in search of a job. Consequently, selection bias was likely present in this study at the analysis unit (inhabitants of households). Another important limitation was the information bias as a result of obtaining group information of participants enrolled in each household through the family head, such as cultural practices shared by all household inhabitants. However, this study considered that some habits and perceptions are widespread in households depending on the region or area where people have grown or lived; for instance, the risk perceptions related to infectious diseases, self-care measures, or land use. For the seroincidence analysis, the main limitation was the losses to follow-up. Selection bias was likely introduced in the estimation of the overall cumulative seroincidence, serogroups seroincidence, and associated factors because the majority of the losses were males. Another potential limitation was the sensitivity and specificity of the microagglutination test to detect the seroincident and seroprevalent cases, especially in seropositive individuals who had titers equal to 1:50. Nevertheless, the sensitivity and specificity of the test could be increased when diverse serovars are included, especially those circulating in the same area. In this study, 32 *Leptospira* strains representing several species and serogroups (Table 1) were included, and a random selection of 20% of the total human serum samples collected at T0 was used to select the final panel of 16 *Leptospira* strains tested in this study. In addition, the classification of most probable infecting serogroup (*L. interrogans* and other *Leptospira* species serogroups) by MAT is not perfect and it could result in misclassification bias. Finally, information biases are also a concern in the followed-up participants, specifically, memory biases respecting changing exposures such as episodes of fever, sporadic outdoor occupations, and contact with synanthropic, wild, or domestic mammals, among other variables.

## CONCLUSIONS

The current study aimed to understand the potential public health problem of *Leptospira* transmission in the rural areas of Urabá, Colombia, where fatal cases of leptospirosis have occurred. The estimated *Leptospira* seroprevalence was 27.81%, with infections being predominantly caused by serogroups of *L. interrogans* (65.66%). The overall seroincidence of *Leptospira* was 14.60%, and a contrast between seroprevalence results with a very similar distribution between *L. interrogans* and other *Leptospira* species serogroups was evident. Also, we identified important risk factors for *Leptospira* infection, which indicates that although different activities and characteristics are related to different species of *Leptospira*, there is a major risk of infection caused by different species circulating in the area. The sequential binary and multinomial regression analysis of this study allowed us to obtain the estimated risk and protective factors associated with *Leptospira* infection (seroprevalent and seroincident cases) in rural areas of the Urabá region. The characteristics of the cases of *Leptospira* infection identified in this study agreed with the cases reported in the Colombian surveillance system for *Leptospira*^48^. Also, this rural area seems to have different species of *Leptospira* circulating, from other areas such as Salvador Bahia, Brazil, which is more urban area and where the rat has a major role in transmission. But this study is consistent with other works showing that transmission in rural areas is also dependent on wild animals and environmental conditions favoring the risk of infection, making disease control difficult and highlights the importance of this study, which identify factors associated to *Leptospira* infection. The longterm goal of this work is to strengthen the surveillance system by identifying areas at risk for *Leptospira* transmission. This, in turn, will aid targeted efforts to provide appropriate diagnosis and timely treatment and ultimately reduce the case fatality rate associated with the disease. Finally, this study will help to prioritize and develop strategies to prevent incident cases in this area and will help to strengthen educational strategies to prevent the transmission of the disease in the communities from rural areas of Colombia.

## Data Availability

All data produced in the present work are contained in the manuscript

## ACKNOWLEDGEMENTS

We are thankful to the research groups Salud y Ambiente and Epidemiología of Facultad Nacional de Salud Pública from Universidad de Antioquia and the Laboratory of Epidemiology of Microbial Diseases from Yale School of Public Health for their logistic support to conduct this study. We also recognize the help from the communities of Alto de Mulatos and Las Changas from the Urabá region. We express our gratitude to the Fogarty International Center for the help and advice obtained through the Research Training Program on the Impact of Zoonotic and Vector-borne Viruses, Rickettsiae, and Leptospira in Acute Undifferentiated Febrile Illnesses (5D43TW010331-05), NIH through the Naturally Acquired and Vaccine-Mediated Immunity to Leptospirosis grant (R01AI121207), and Departamento Administrativo de Ciencia, Tecnología e Innovación (Colciencias) (award: 111565741009). The funders had no role in the study design, data collection and analysis, decision to publish, or preparation of the manuscript.

