## Supplemental table 1 for "*Leptospira* infection in rural areas of Urabá region, Colombia: a prospective study"

**Table S1.** Descriptive, bivariate and multivariate analysis of seroprevalent cases of *Leptospira* (mixed-effect binary logistic regression model)

| **Variables** | **Seronegative** | ***Leptospira* seropositive** | | |
| --- | --- | --- | --- | --- |
|  | **n = 431 (%)** | **n=166 (%)** | **OR_crude_ (95%CI)** | **OR_adjust_ (95%CI)** |
| **Individuals** | | | | |
| Outdoor occupation | 101 (23.43) | 71 (42.77) | 2.55 (1.91-3.42) | 1.56 (1.05-2.31) |
| Male gender | 146 (33.87) | 87 (52.41) | 2.24 (1.69-2.96) | 2.01 (1.42-2.84) |
| Age (years): median (IQR) | 29.47 (14.35-45.66) | 31.43 (18.94-48.72) | 1.01 (1.00-1.02) |  |
| Age, years (≤1-15) | 120 (25.84) | 30 (18.07) | 1.00 | 1.00 |
| Age, years (>15-29) | 99 (22.97) | 50 (30.12) | 2.23 (1.48-3.37) | 2.30 (1.47-3.60) |
| Age, years (>29-46) | 110 (25.52) | 39 (23.49) | 1.47 (0.96-2.25) | 1.50 (0.92-2.42) |
| Age, years (>46) | 102 (23.67) | 47 (28.31) | 1.99 (1.32-3.04) | 1.80 (1.11-2.92) |
| Ethnicity (Afro-descendant and Amerindians) | 23 (5.36) | 14 (8.43) | 1.74 (1.00-3.03) |  |
| History of fever | 246 (57.08) | 104 (62.65) | 1.29 (0.97-1.71) | 1.42 (1.05-1.91) |
| Time of residence in the area; Median (QIR) | 10.00 (5.00-18.00) | 12.00 (7.00-20.00) | 1.01 (0.99-1.02) |  |
| **Household characteristics** | | | | |
| **Roof material** |  |  |  |  |
| Zinc | 304 (70.53) | 103 (17.25) | 0.66 (0.50-0.89) |  |
| Vegetal (palm) | 204 (47.33) | 88 (53.01) | 1.25 (0.95-1.64) |  |
| Cement | 5 (1.16) | 1 (0.60) | 0.52 (0.10-2.75) |  |
| Tile | 16 (3.71) | 7 (4.22) | 1.12 (0.54-2.30) |  |
| Wood | 26 (6.03) | 10 (6.02) | 0.95 (0.53-1.72) |  |
| **Floor material** |  |  |  |  |
| Dirt soil | 309 (71.69) | 132 (79.52) | 1.51 (1.08-2.09) | 1.62 (1.14-2.31) |
| Cement | 187 (43.39) | 66 (39.76) | 0.85 (0.64-1.12) |  |
| Wood | 34 (7.89) | 9 (5.42) | 0.73 (0.41-1.31) |  |
| Tile | 22 (5.10) | 5 (3.01) | 0.56 (0.27-1.19) |  |
| **Wall material** |  |  |  |  |
| Wood | 387 (89.79) | 153 (92.17) | 1.89 (0.84-2.17) |  |
| Brick wall | 130 (30.16) | 45 (27.11) | 0.87 (0.65-1.18) |  |
| **Public services in household** | | | | |
| Presence of aqueduct | 258 (59.86) | 85 (51.20) | 0.69 (0.52-0.91) |  |
| Waste disposal | 249 (57.77) | 81 (48.80) | 0.69 (0.52-0.91) |  |
| Presence of sewage | 62 (14.39) | 20 (12.05) | 0.82 (0.54-1.24) |  |
| Presence of latrine | 230 (53.36) | 77 (46.39) | 0.75 (0.57-0.99) |  |
| Presence of pit latrine | 42 (9.74) | 24 (14.46) | 1.59 (1.03-2.44) |  |
| **Household location** | | | | |
| Urban center | 219 (50.81) | 74 (44.58) | 0.77 (0.58-1.01) |  |
| Rural | 212 (49.19) | 92 (55.42) | 1.00 |  |
| **Household proximity** | | | | |
| Very near | 99 (22.97) | 39 (23.49) | 1.00 |  |
| Near | 147 (34.11) | 50 (30.12) | 0.91 (0.63-1.31) |  |
| Scattered | 113 (26.22) | 34 (20.48) | 0.81 (0.54-1.21) |  |
| Very scattered | 72 (16.71) | 43 (25.90) | 1.58 (1.05-2.38) |  |
| **Peri domiciliary area characteristics** | | | | |
| **Vegetation** |  |  |  |  |
| Bushes | 396 (91.88) | 156 (93.98) | 1.40 (0.82-2.39) |  |
| Trees | 380 (88.17) | 152 (91.57) | 1.54 (0.97-2.44) |  |
| Grasses | 228 (52.90) | 86 (51.81) | 0.97 (0.74-1.28) |  |
| Corn culture | 12 (2.78) | 6 (3.61) | 1.42 (0.63-3.20) |  |
| Cassava culture | 23 (5.34) | 13 (7.83) | 1.54 (0.89-2.66) |  |
| Tomato culture | 13 (3.02) | 3 (1.81) | 0.59 (0.22-1.59) |  |
| **Presence of wild animals in peri domiciliary area** | | | | |
| Opossum | 196 (46.23) | 92 (55.42) | 1.46 (1.11-1.93) | 1.34 (1.003-1.80) |
| Wild animals | 122 (29.33) | 52 (32.70) | 1.23 (0.90-1.67) |  |
| **Presence of domestic animals in intra or peri domiciliary area** | | | | |
| Canines | 251 (58.24) | 100 (60.24) | 1.11 (0.84-1.46) |  |
| Felines | 257 (59.63) | 107 (64.46) | 1.19 (0.90-1.58) |  |
| Poultry | 301 (69.84) | 114 (68.67) | 0.95 (0.70-1.29) |  |
| Turkeys | 113 (26.22) | 50 (30.12) | 1.22 (0.90-1.66) |  |
| Porcine | 201 (46.64) | 87 (52.41) | 1.32 (1.00-1.74) |  |
| Horses | 111 (25.75) | 41 (24.70) | 0.98 (0.71-1.35) |  |
| Donkeys | 118 (27.38) | 52 (31.33) | 1.25 (0.92-1.70) |  |
| Mules | 62 (14.39) | 26 (15.66) | 1.13 (0.77-1.66) |  |
| **Animal husbandry purpose** | | | | |
| Hunting canines | 15 (3.48) | 6 (3.61) | 1.02 (0.47-2.12) |  |
| Canines (companionship) | 240 (55.68) | 95 (57.23) | 1.09 (0.83-1.44) |  |
| Poultry (eggs) | 247 (57.31) | 87 (52.41) | 0.82 (0.62-1.09) |  |
| Poultry (meat) | 173 (40.14) | 69 (41.57) | 1.03 (0.78-1.36) |  |
| Poultry (companionship) | 22 (5.10) | 13 (7.83) | 1.63 (0.94-2.85) |  |
| Porcine (Fattening) | 181 (42.00) | 81 (48.80) | 1.35 (1.02-1.77) |  |
| Porcine (piglet) | 78 (18.10) | 44 (26.51) | 1.70 (1.23-2.37) | 1.45 (1.02-2.07) |
| Porcine (companionship) | 6 (1.39) | 3 (1.81) | 1.59 (0.55-4.56) |  |
| Turkeys (eggs) | 70 (16.24) | 31 (18.67) | 1.20 (0.83-1.73) |  |
| Turkeys (meat) | 98 (22.74) | 39 (23.49) | 1.08 (0.78-1.49) |  |
| Turkeys (companionship) | 10 (2.32) | 9 (5.42) | 2.45 (1.16-5.17) |  |
| Packhorses | 75 (17.63) | 28 (16.87) | 0.97 (0.68-1.40) |  |
| Horses (cowboy) | 45 (10.44) | 16 (9.64) | 0.93 (0.58-1.48) |  |
| Horses (companionship) | 7 (1.62) | 3 (1.81) | 1.09 (0.36-3.33) |  |
| Pack donkeys | 114 (26.45) | 51 (30.72) | 1.27 (0.93-1.72) |  |
| Donkeys (companionship) | 4 (0.93) | 1 (0.60) | 0.73 (0.12-4.50) |  |
| Pack mules | 57 (13.23) | 26 (15.66) | 1.23 (0.83-1.82) |  |
| Mules (cowboy) | 10 (2.32) | 0 (0) | NS |  |
| **Synanthropic mammals** | | | | |
| Rats | 340 (78.89) | 141 (84.94) | 1.60 (1.11-2.31) |  |
| **Practices common among families in study area** | | | | |
| Forest fragmentation and deforestation | 267 (61.95) | 111 (66.87) | 1.30 (0.98-1.72) |  |
| Use of any rodent’s elimination measure | 249 (57.77) | 104 (62.65) | 1.23 (0.92-1.63) |  |
