## Supplemental table 2 for "*Leptospira* infection in rural areas of Urabá region, Colombia: a prospective study"

**Table S2.** Descriptive and bivariate analysis of seroprevalent cases of *Leptospira* among different serogroups (mixed-effects multinomial logistic regression model)

| **Variables** | **Seronegative** | ***L. interrogans* serogroup** | | **Others *Leptospira* species serogroups** | | |
| --- | --- | --- | --- | --- | --- | --- |
|  | **n = 431 (%)** | **n=109 (%)** | **OR_crude_ (95%CI)** | **n=57 (%)** | | **OR_crude_ (95%CI)** |
| **Individuals** | | | | | | |
| Outdoor occupation | 101 (23.43) | 57 (52.29) | 3.72 (2.66-5.20) | 14 (24.56) | | 1.12 (0.68-1.83) |
| Male gender | 146 (33.87) | 64 (58.72) | 2.99 (2.14-4.17) | 23 (40.35) | | 1.33 (0.86-2.05) |
| Age (years): median (IQR) | 29.47 (14.35-45.66) | 33.23 (19.67-49.25) |  | 28.42 (16.76-44.66) | |  |
| Age, years (1-15) | 120 (25.84) | 16 (14.68) | 1.00 | 14 (24.56) | | 1.00 |
| Age, years (>15-29) | 99 (22.97) | 33 (30.28) | 2.77 (1.65-4.63) | 17 (29.82) | | 1.61 (0.89-2.90) |
| Age, years (>29-46) | 110 (25.52) | 27 (24.77) | 1.95 (1.15-3.30) | 12 (21.05) | | 0.92 (0.48-1.74) |
| Age, years (>46) | 102 (23.67) | 33 (30.28) | 2.61 (1.56-4.37) | 14 (24.56) | | 1.28 (0.70-2.34) |
| Ethnicity (Afro-descendant and Amerindians) | 23 (5.36) | 12 (11.01) | 2.28 (1.26-4.13) | 2 (3.51) | | 0.68 (0.21-2.23) |
| History of fever | 246 (57.08) | 67 (61.47) | 1.25 (0.89-1.74) | 37 (64.91) | | 1.41 (0.90-2.20) |
| Time of residence in the area; Median (QIR) | 10.00 (5.00-18.00) | 13.00 (7.00-20.00) | 1.01 (0.993-1.019) | 12 (7-24) | | 1.013 (0.997-1.029) |
| **Household characteristics** | | | | | | |
| **Roof material** |  |  |  |  |  | |
| Zinc | 304 (70.53) | 66 (60.55) | 0.63 (0.45-0.88) | 37 (64.91) | 0.74 (0.47-1.16) | |
| Vegetal (palm) | 204 (47.33) | 60 (55.05) | 1.33 (0.96-1.83) | 28 (49.12) | 1.01 (0.72-1.68) | |
| Cement | 5 (1.16) | 1 (0.92) | 0.90 (0.17-4.96) | 0 (0) | NS | |
| Tile | 16 (3.71) | 5 (4.59) | 1.29 (0.55-3.02) | 2 (3.51) | 0.99 (0.31-3.19) | |
| Wood | 26 (6.03) | 6 (5.50) | 0.86 (0.41-1.78) | 4 (7.02) | 1.15 (0.49-2.70) | |
| **Floor material** |  |  |  |  |  | |
| Dirt soil | 309 (71.69) | 89 (81.65) | 1.71 (1.13-2.59) | 43 (75.44) | 1.19 (0.73-1.95) | |
| Cement | 187 (43.39) | 41 (37.61) | 0.80 (0.55-1.15) | 25 (43.86) | 0.99 (0.65-1.53) | |
| Wood | 34 (7.89) | 4 (3.67) | 0.50 (0.22-1.12) | 5 (8.77) | 1.25 (0.59-2.65) | |
| Tile | 22 (5.10) | 4 (3.67) | 0.71 (0.31-1.61) | 1 (1.75) | 0.31 (0.07-1.50) | |
| **Wall material** |  |  |  |  |  | |
| Wood | 387 (89.79) | 101 (92.66) | 1.36 (0.77-2.42) | 52 (91.23) | 1.27 (0.61-2.63) | |
| Brick wall | 130 (30.16) | 27 (24.77) | 0.81 (0.55-1.18) | 18 (31.58) | 1.05 (0.67-1.66) | |
| **Public services in household** | | | | | | |
| Presence of aqueduct | 258 (59.86) | 54 (49.54) | 0.61 (0.47-0.90) | 31 (54.39) | 0.76 (0.50-1.18) | |
| Waste disposal | 249 (57.77) | 52 (47.71) | 0.67 (0.48-0.93) | 29 (4.86) | 0.73 (0.47-1.12) | |
| Presence of sewage | 62 (14.39) | 10 (9.17) | 0.60 (0.35-1.04) | 10 (17.54) | 1.29 (0.73-2.27) | |
| Presence of latrine | 230 (53.36) | 50 (45.87) | 0.72 (0.50-1.018) | 27 (47.37) | 0.75 (0.49-1.15) | |
| Presence of pit latrine | 42 (9.74) | 14 (12.84) | 1.39 (0.80-2.39) | 10 (17.54) | 2.06 (1.13-3.75) | |
| **Household location** | | | | | | |
| Urban center | 219 (50.81) | 46 (42.20) | 0.69 (0.50-0.96) | 28 (49.12) | 0.93 (0.61-1.42) | |
| Rural | 212 (49.19) | 63 (57.80) | 1.00 | 29 (50.88) | 1.00 | |
| **Household proximity** | | | | | | |
| Very near | 99 (22.97) | 21 (19.27) | 1.00 | 18 (31.58) | 1.00 | |
| Near | 147 (34.11) | 32 (29.36) | 1.06 (0.42-1.23) | 18 (31.58) | 0.72 (0.42-1.23) | |
| Scattered | 113 (26.22) | 23 (21.10) | 0.99 (0.61-1.63) | 11 (19.30) | 0.57 (0.31-1.06) | |
| Very scattered | 72 (16.71) | 33 (30.28) | 2.18 (1.34-3.53) | 10 (8.70) | 0.82 (0.43-1.59) | |
| **Peri domiciliary area characteristics** | | | | | | |
| **Vegetation** |  |  |  |  |  | |
| Bushes | 396 (91.88) | 102 (93.58) | 1.35 (0.73-2.51) | 54 (94.74) | 1.74 (0.70-4.35) | |
| Trees | 380 (88.17) | 99 (90.83) | 1.39 (0.81-2.39) | 53 (92.98) | 1.80 (0.82-3.93) | |
| Grasses | 228 (52.90) | 55 (50.46) | 0.91 (0.65-1.27) | 31 (54.39) | 1.16 (0.76-1.78) | |
| Corn culture | 12 (2.78) | 4 (3.67) | 1.30 (0.49-3.45) | 2 (3.51) | 1.41 (0.41-4.80) | |
| Cassava culture | 23 (5.34) | 10 (9.17) | 1.89 (1.04-3.42) | 3 (5.26) | 0.86 (0.31-2.39) | |
| Tomato culture | 13 (3.02) | 3 (2.75) | 1.06 (0.38-3.02) | 0 (0) | NS | |
| **Presence of wild animals in peri domiciliary area** | | | | | | |
| Opossum | 196 (46.23) | 66 (60.55) | 1.76 (1.27-2.43) | 26 (45.61) | 1.03 (0.67-1.58) | |
| Wild animals | 122 (29.33) | 38 (35.85) | 1.39 (0.98-1.98) | 14 (26.42) | 0.92 (0.55-1.53) | |
| **Presence of domestic animals in intra or peri domiciliary area** | | | | | | |
| Canines | 252 (58.24) | 63 (57.80) | 0.92 (0.65-1.32) | 37 (64.91) | 1.38 (0.88-2.14) | |
| Felines | 257 (59.63) | 70 (64.22) | 1.12 (0.79-1.60) | 37 (64.91) | 1.22 (0.79-1.91) | |
| Poultry | 301 (69.84) | 71 (65.14) | 0.75 (0.53-1.08) | 43 (75.44) | 1.37 (0.84-2.23) | |
| Turkeys | 113 (26.22) | 32 (29.36) | 1.08 (0.71-1.64) | 18 (31.58) | 1.34 (0.84-2.13) | |
| Porcine | 201 (46.64) | 60 (55.05) | 1.49 (1.08-2.06) | 27 (47.37) | 1.05(0.68-1.60) | |
| Horses | 111 (25.75) | 26 (23.85) | 0.94 (0.64-1.38) | 15 (26.32) | 1.06 (0.65-1.73) | |
| Donkeys | 118 (27.38) | 33 (30.28) | 1.09 (0.73-1.64) | 19 (33.33) | 1.44 (0.91-2.27) | |
| Mules | 62 (14.39) | 14 (12.84) | 0.93 (0.57-1.50) | 12 (21.05) | 1.58 (0.92-2.72) | |
| **Animal husbandry purpose** | | | | | | |
| Hunting canines | 15 (3.48) | 2 (1.83) | 0.41 (0.12-1.41) | 4 (7.02) | 2.08 (0.84-5.16) | |
| Canines (companionship) | 240 (55.68) | 61 (55.96) | 0.98 (0.70-1.37) | 34 (59.65) | 1.23 (0.80-1.90) | |
| Poultry (eggs) | 247 (57.31) | 56 (51.38) | 0.77 (0.55-1.07) | 31 (54.39) | 0.89 (0.58-1.36) | |
| Poultry (meat) | 173 (40.14) | 46 (42.20) | 0.99 (0.70-1.41) | 23 (40.35) | 1.00 (0.65-1.55) | |
| Poultry (companionship) | 22 (5.10) | 6 (5.50) | 0.96 (0.45-2.06) | 7 (12.28) | 2.87 (1.43-5.73) | |
| Porcine (Fattening) | 181 (42.00) | 55 (50.46) | 1.44 (1.04-1.98) | 26 (45.61) | 1.19 (0.77-1.82) | |
| Porcine (piglet) | 78 (18.10) | 33 (30.28) | 2.05 (1.42-2.96) | 2 (19.30) | 1.13 (0.65-1.95) | |
| Porcine (companionship) | 6 (1.39) | 2 (1.83) | 1.44 (0.42-4.99) | 1 (1.75) | 1.69 (0.36-7.85) | |
| Turkeys (eggs) | 70 (16.24) | 22 (20.18) | 1.26 (0.79-2.00) | 9 (15.79) | 1.00 (0.56-1.81) | |
| Turkeys (meat) | 98 (22.74) | 26 (23.85) | 1.00 (0.66-1.53) | 13 (22.81) | 1.07 (0.65-1.78) | |
| Turkeys (companionship) | 10 (2.32) | 4 (3.67) | 1.39 (0.51-3.79) | 5 (8.77) | 4.37 (1.80-10.61) | |
| Packhorses | 75 (17.63) | 18 (16.51) | 0.97 (0.63-1.50) | 10 (17.54) | 0.98 (0.56-1.73) | |
| Horses (cowboy) | 45 (10.44) | 11 (10.09) | 0.96 (0.55-1.66) | 5 (8.77) | 0.85 (0.40-1.81) | |
| Horses (companionship) | 7 (1.62) | 2 (1.83) | 0.92 (0.23-3.58) | 1 (1.75) | 1.13 (0.21-6.15) | |
| Pack donkeys | 114 (26.45) | 32 (29.36) | 1.08 (0.72-1.64) | 19 (33.33) | 1.50 (0.95-2.36) | |
| Donkeys (companionship) | 4 (0.93) | 1 (0.92) | 1.13 (0.18-7.39) | 0 (0) | NS | |
| Pack mules | 57 (13.23) | 14 (12.84) | 1.01 (0.62-1.64) | 12 (21.05) | 1.73 (1.00-2.98) | |
| Mules (cowboy) | 10 (2.32) | 0 (0) | NS | 0 (0) | NS | |
| **Synanthropic mammals** | | | | | | |
| Rats | 340 (78.89) | 92 (84.40) | 1.56 (1.01-2.40) | 49 (85.96) | 1.74 (0.96-3.17) | |
| **Practices common among families in study area** | | | | | | |
| Forest fragmentation and deforestation | 267 (61.95) | 69 (63.30) | 1.08 (0.76-1.52) | 42 (73.68) | 1.87 (1.17-2.98) | |
| Use of any rodent’s elimination measure | 249 (57.77) | 73 (66.97) | 1.52 (1.08-2.14) | 31 (54.39) | 0.84 (0.55-1.29) | |
