## Supplemental table 3 for "*Leptospira* infection in rural areas of Urabá region, Colombia: a prospective study"

**Table S3.** Characteristics of individuals included in the study and lost to the follow-up.

| **Baseline characteristics** | **Follow-up at T12 Alto de Mulatos n=120 (%)** | **Lost to follow-up at T12 Alto de Mulatos n=135 (%)** | **Follow-up at T12 Las Changas n=154 (%)** | **Lost to follow-up at T12 Las Changas n=188 (%)** |
| --- | --- | --- | --- | --- |
| Age in years (Median, IQR) | 32.70 (15.67-47.34) | 23.76 (12.93-39.94) | 35.65 (19.58-53.37) | 28.03 (13.85-44.57) |
| Male gender | 35 (29.17) | 73 (54.07) | 48 (31.17) | 77 (40.96) |
| Outdoor occupation (last year) | 14 (11.67) | 30 (22.22) | 55 (35.71) | 54 (28.72) |
| Presence of rodents (peri domiciliary area) | 100 (83.33) | 122 (90.37) | 112 (72.73) | 147 (78.19) |
| Presence of opossums (peri domiciliary area) | 64 (53.33) | 44 (32.84) | 87 (56.86) | 93 (50.82) |
| Presence of pit latrine | 30 (25.00) | 36 (26.00) | 0 (0.0) | 0 (0.0) |
| Presence of dirty soil floor | 66 (55.00) | 97 (71.85) | 121 (78.57) | 157 (83.51) |
