## Supplemental table 4 for "*Leptospira* infection in rural areas of Urabá region, Colombia: a prospective study"

**Table S4.** Descriptive, bivariate and multivariate analysis of seroincident cases of *Leptospira* (mixed-effect binary logistic regression model)

| **Variables** | **Seronegative** | ***Leptospira* seropositive** | | |
| --- | --- | --- | --- | --- |
|  | **n = 234 (%)** | **n=40 (%)** | **RR_crude_ (95%CI)** | **RR_adjust_ (95%CI)^a^** |
| **Individuals** | | | | |
| Outdoor occupation | 55 (23.50) | 14 (35.00) | 1.57 (1.04-2.30) | 2.66 (1.46-4.68) |
| Male gender | 72 (30.77) | 11 (27.50) | 0.79 (0.49-1.23) |  |
| Age (years): median (IQR) | 34.18 (18.11 – 50.87) | 40.30 (20.46-55.34) | 1.01 (1.00-1.02) |  |
| Ethnicity (Afro-descendant and Amerindians) | 15 (6.41) | 2 (5.13) | 0.74 (0.26-1.76) |  |
| History of fever in last 12 months | 13 (5.56) | 1 (2.50) | 0.46 (0.12-1.55) |  |
| Time of residence in the area; Median (QIR) | 13.00 (7.00 -23.00) | 14.00 (8.00 -21.00) | 1.00 (0.99-1.02) |  |
| **Household characteristics** | | | | |
| **Roof material** |  |  |  |  |
| Zinc | 155 (66.24) | 26 (65.00) | 0.97 (0.63-1.45) |  |
| Vegetal (palm) | 114 (48.72) | 22 (55.00) | 1.23 (0.82-1.80) |  |
| Wood | 20 (8.55) | 4 (16.67) | 1.20 (0.57-2.27) |  |
| **Floor material** |  |  |  |  |
| Dirt soil | 155 (66.24) | 32 (80.00) | 1.82 (1.11-2.85) | 1.77 (1.01-3.06) |
| Cement | 102 (43.59) | 10 (25.00) | 0.46 (0.28-0.73) |  |
| Wood | 22 (9.40) | 1 (2.50) | 0.34 (0.09-1.17) |  |
| Tile | 14 (5.98) | 2 (5.00) | 0.93 (0.37-2.08) |  |
| **Wall material** |  |  |  |  |
| Wood | 209 (89.32) | 37 (92.50) | 1.24 (0.61-2.34) |  |
| Brick wall | 77 (32.91) | 6 (15.00) | 0.46 (0.26-0.77) |  |
| **Public services in household** | | | | |
| Presence of aqueduct | 129 (55.13) | 17 (42.50) | 0.69(1.97-1.03) |  |
| Waste disposal | 126 (53.85) | 16 (40.00) | 0.67 (0.44-1.00) |  |
| Presence of sewage | 30 (12.82) | 4 (10.00) | 0.83 (0.42-1.54) |  |
| Presence of latrine | 113 (48.29) | 17 (42.50) | 0.87 (0.57-1.28) |  |
| Presence of pit latrine | 23 (9.83) | 7 (17.50) | 1.57 (0.88-2.57) |  |
| **Household location** | | | | |
| Urban center | 111 (47.44) | 16 (40.00) | 0.85 (0.56-1.24) |  |
| Rural | 123 (52.56) | 24 (60.00) | 1.00 |  |
| **Household proximity** | | | | |
| Very near | 56 (23.93) | 8 (20.00) | 1.00 |  |
| Near | 71 (30.34) | 10 (25.00) | 0.95(0.52-1.66) |  |
| Scattered | 64 (27.35) | 10 (25.00) | 1.04 (0.57-1.82) |  |
| Very scattered | 43 (18.38) | 12 (30.00) | 1.60 (0.91-2.62) |  |
| **Peri domiciliary area characteristics** | | | | |
| **Vegetation** |  |  |  |  |
| Bushes | 219 (93.59) | 35 (87.50) | 0.51 (0.27-0.92) |  |
| Trees | 216 (92.31) | 38 (95.00) | 1.43 (0.59-3.09) |  |
| Grasses | 123 (52.56) | 23 (57.50) | 1.17 (0.78-1.71) |  |
| Corn culture | 2 (0.85) | 2 (5.00) | 3.72 (1.42-6.02) | 8.33 (2.21-19.65) |
| Cassava culture | 15 (6.41) | 6 (15.00) | 2.18 (1.25-3.43) |  |
| Tomato culture | 4 (1.71) | 1 (2.50) | 1.45 (0.38-3.83) |  |
| **Presence of wild animals in peri domiciliary area** | | | | |
| Opossum | 134 (57.26) | 17 (43.59) | 0.60 (0.39-0.91) |  |
| Wild animals | 68 (30.09) | 12 (30.77) | 1.05(0.66-1.60) |  |
| **Presence of domestic animals in intra or peri domiciliary area** | | | | |
| Canines | 135 (57.69) | 26 (65.00) | 1.26 (0.83-1.86) |  |
| Felines | 144 (61.54) | 26 (65.00) | 1.09 (0.71-1.62) |  |
| Poultry | 171 (73.08) | 26 (65.00) | 0.72 (0.46-1.09) |  |
| Turkeys | 55 (23.50) | 11 (27.50) | 1.20 (0.75-1.85) |  |
| Porcine | 115 (49.15) | 20 (50.00) | 1.04 (0.69-1.53) |  |
| Horses | 69 (29.49) | 12 (30.00) | 1.01 (0.64-1.53) |  |
| Donkeys | 74 (31.62) | 15 (37.50) | 1.21 (0.79-1.78) |  |
| Mules | 32 (13.68) | 2 (5.00) | 0.37 (0.14-0.93) |  |
| **Animal husbandry purpose** | | | | |
| Hunting canines | 7 (2.99) | 3 (7.50) | 2.25 (1.05-3.98) | 5.16 (1.83-10.74) |
| Canines (companionship) | 130 (55.56) | 24 (60.00) | 1.12 (0.74-1.65) |  |
| Poultry (eggs) | 136 (58.12) | 21 (52.50) | 0.82 (0.54-1.21) |  |
| Poultry (meat) | 96 (41.03) | 14 (35.00) | 0.82 (0.53-1.23) |  |
| Poultry (companionship) | 8 (3.42) | 3 (7.50) | 2.14 (1.01-3.78) |  |
| Porcine (Fattening) | 99 (42.31) | 17 (42.50) | 1.00 (0.66-1.48) |  |
| Porcine (piglet) | 52 (22.22) | 12 (30.00) | 1.50 (0.97-2.22) |  |
| Porcine (companionship) | 3 (1.28) | 1 (2.50) | 1.63(0.39-4.34) |  |
| Turkeys (eggs) | 40 (17.09) | 8 (20.00) | 1.14 (0.67-1.85) |  |
| Turkeys (meat) | 47 (20.09) | 8 (20.00) | 1.03 (0.61-1.67) |  |
| Turkeys (companionship) | 2 (0.85) | 1 (2.50) | 2.51 (0.64-5.41) |  |
| Packhorses | 47 (20.09) | 10 (25.00) | 1.27 (0.80-1.96) |  |
| Horses (cowboy) | 27 (11.54) | 3 (7.50) | 0.68 (0.31-1.40) |  |
| Horses (companionship) | 7 (2.99) | 1 (2.50) | 0.92 (0.23-2.81) |  |
| Pack donkeys | 73 (31.20) | 15 (37.50) | 1.22 (0.80-1.81) |  |
| Pack mules | 31 (13.25) | 2 (5.00) | 0.38 (0.14-0.96) |  |
| **Synanthropic mammals** | | | | |
| Rats | 178 (76.07) | 34 (85.00) | 1.57 (0.91-2.59) | 2.00 (1.03-3.76) |
| **Practices common among families in study area** | | | | |
| Forest fragmentation and deforestation | 144 (61.54) | 26 (65.00) | 1.12 (0.74-1.67) |  |
| Use of any rodent’s elimination measure | 131 (55.98) | 21 (52.50) | 0.94 (0.62-1.39) |  |

^a^ Adjusted by gender and age
