## Supplemental table 5 for "*Leptospira* infection in rural areas of Urabá region, Colombia: a prospective study"

**Table S4.** Descriptive and bivariate analysis of seroincident cases of *Leptospira* among different serogroups (mixed-effects multinomial logistic regression model)

| **Variables** | **Seronegative** | ***L. interrogans* serogroup** | | | **Others *Leptospira* species serogroups** | |
| --- | --- | --- | --- | --- | --- | --- |
|  | **n = 234 (%)** | **n=23 (%)** | **RR_crude_ (95%CI)** | **n=17 (%)** | | **RR_crude_ (95%CI)** |
| **Individuals** |  |  |  |  | |  |
| Outdoor occupation | 55 (23.50) | 9 (39.13) | 1.85 (1.07-3.06) | 5 (29.41) | | 1.34 (0.65-2.65) |
| Male gender | 72 (30.77) | 7 (30.43) | 0.87 (0.47-1.55) | 4 (23.53) | | 0.64 (0.29-1.40) |
| Age (years); Median (QIR) | 34.18 (18.11 – 50.87) | 47.46 (24.76 – 58.42) | 1.02 (1.00-1.03) | 29.99 (18.19 – 49.28) | | 0.99 (0.98-1.01) |
| Ethnicity (Afro-descendant and Amerindians) | 15 (6.41) | 1 (4.35) | 0.66 (0.41-2.46) | 1 (6.25) | | 0.72 (0.13-3.55) |
| History of fever in last 12 months | 13 (5.56) | 1 (4.35) | 0.75 (0.19-2.54) | 0 (0.00) | | NS |
| Time of residence in the area; Median (QIR) | 13.00 (7.00 -23.00) | 14.00 (9.00 – 19.00) | 1.00 (0.98-1.02) | 11.00 (7.00 -25.00) | | 1.01 (0.98-1.03) |
| **Household characteristics** | | | | | | |
| **Roof material** |  |  |  |  | |  |
| Zinc | 155 (66.24) | 15 (65.22) | 1.00 (0.56-1.73) | 11 (64.71) | | 0.95 (0.42-2.09) |
| Vegetal (palm) | 114 (48.72) | 13 (56.52) | 1.32 (0.77-2.21) | 9 (52.94) | | 1.14 (0.42-2.51) |
| Wood | 20 (8.55) | 2 (8.70) | 1.04 (0.36-2.68) | 2 (11.76) | | 1.44 (0.48-3.91) |
| **Floor material** |  |  |  |  | |  |
| Dirt soil | 155 (66.24) | 19 (82.61) | 2.21 (1.10-4.19) | 13 (76.47) | | 1.53 (0.71-3.18) |
| Cement | 102 (43.59) | 6 (26.09) | 0.47 (0.25-0.87) | 4 (23.53) | | 0.38 (0.17-0.32) |
| Wood | 22 (9.40) | 1 (4.35) | 0.54 (0.14-1.91) | 0 (0.00) | | NS |
| Tile | 14 (5.98) | 1 (4.35) | 0.87 (0.25-2.68) | 1 (5.88) | | 0.98 (0.25-3.65) |
| **Wall material** |  |  |  |  | |  |
| Wood | 209 (89.32) | 21 (91.30) | 1.11 (0.93-2.53) | 16 (94.50) | | 1.55 (0.42-5.13) |
| Brick wall | 77 (32.91) | 4 (17.13) | 0.52 (0.26-1.00) | 2 (11.76) | | 0.32 (0.12-0.83) |
| **Public services in household** | | | | | | |
| Presence of aqueduct | 129 (55.13) | 12 (52.17) | 0.94 (0.54-1.59) | 5 (29.41) | | 0.40 (0.19-0.80) |
| Waste disposal | 126 (53.85) | 11 (47.83) | 0.87 (0.49-1.49) | 5 (29.41) | | 0.53 (0.29-0.89) |
| Presence of sewage | 30 (12.82) | 2 (8.70) | 0.69 (0.26-1.71) | 2 (11.76) | | 2.58 (0.36-2.85) |
| Presence of latrine | 113 (48.29) | 11 (47.83) | 1.09 (0.63-1.83) | 6 (35.29) | | 0.60 (0.30-1.17) |
| Presence of pit latrine | 23 (9.83) | 4 (17.39) | 1.62 (0.75-3.20) | 3 (17.65) | | 1.64 (0.63-3.87) |
| **Household location** | | | | | | |
| Urban center | 111 (47.44) | 11 (47.83) | 1.11 (0.64-1.87) | 5 (29.41) | | 0.55 (0.28-1.06) |
| Rural | 123 (52.56) | 12 (52.17) | 1.00 | 12 (70.59) | | 1.00 |
| **Household proximity** | | | | | | |
| Very near | 56 (23.93) | 5 (21.74) | 1.00 | 3 (17.65) | | 1.00 |
| Near | 71 (30.34) | 6 (26.09) | 0.96 (0.44-1.98) | 4 (23.53) | | 0.92 (0.33-2.42) |
| Scattered | 64 (27.35) | 6 (26.09) | 0.98 (0.44-2.07) | 4 (23.53) | | 1.15 (0.42-2.93) |
| Very scattered | 43 (18.38) | 6 (26.09) | 1.43 (0.65-2.89) | 6 (35.29) | | 2.06 (0.82-4.72) |
| **Peri domiciliary area characteristics** | | | | | | |
| **Vegetation** |  |  |  |  | |  |
| Bushes | 219 (93.59) | 20 (86.96) | 0.82 (0.57-1.01) | 15 (88.24) | | 0.47 (0.17-1.22) |
| Trees | 216 (92.31) | 21 (91.30) | 0.87 (0.34-2.04) | 17 (100.00) | | NS |
| Grasses | 123 (52.56) | 14 (60.87) | 1.34 (0.77-2.25) | 9 (52.94) | | 0.96 (0.48-1.86) |
| Corn culture | 2 (0.85) | 2 (8.70) | 6.14 (2.32-9.96) | 0 (0.00) | | NS |
| Cassava culture | 15 (6.41) | 3 (13.04) | 1.97 (0.90-4.04) | 3 (17.65) | | 2.95 (1.24-6.18) |
| Tomato culture | 4 (1.71) | 1 (4.35) | 2.36 (0.62-6.26) | 0 (0.00) | | NS |
| **Presence of wild animals in peri-domiciliary area** | | | | | | |
| Opossum | 134 (57.26) | 8 (36.36) | 0.46 (0.25-0.82) | 9 (52.94) | | 0.74 (0.36-1.47) |
| Wild animals | 68 (30.09) | 7 (31.82) | 1.05 (0.57-1.88) | 5 (29.41) | | 0.98 (0.34-266) |
| **Presence of domestic animals in intra or peri-domiciliary area** | | | | | | |
| Canines | 135 (57.69) | 15 (65.22) | 1.25 (0.71-2.12) | 11 (64.71) | | 1.31 (0.63-2.62) |
| Felines | 144 (61.54) | 13 (56.52) | 0.80 (0.46-1.36) | 13 (76.47) | | 1.89 (0.89-3.84) |
| Poultry | 171 (73.08) | 14 (60.87) | 0.09 (0.35-1.07) | 12 (70.59) | | 0.83 (0.39-1.68) |
| Turkeys | 55 (23.50) | 6 (26.09) | 1.09 (0.57-1.99) | 5 (29.41) | | 1.44 (0.69-2.84) |
| Porcine | 115 (49.15) | 10 (43.48) | 0.81 (0.46-1.37) | 10 (58.52) | | 1.54 (0.79-2.90) |
| Horses | 69 (29.49) | 6 (26.09) | 0.81 (0.43-1.49) | 6 (35.29) | | 1.34 (0.67-2.58) |
| Donkeys | 74 (31.62) | 6 (26.09) | 0.73 (0.38-1.34) | 9 (52.54) | | 2.35 (1.23-4.31) |
| Mules | 32 (13.68) | 0 (0.00) | NS | 2 (11.76) | | 0.87 (0.31-2.30) |
| **Animal husbandry purpose** | | | | | | |
| Hunting canines | 7 (2.99) | 0 (0.00) | NS | 3 (17.65) | | 5.71 (2.56-10.31) |
| Canines (companionship) | 130 (55.56) | 15 (65.22) | 1.35 (0.77-2.28) | 9 (52.94) | | 0.85 (0.42-1.67) |
| Poultry (eggs) | 136 (58.12) | 11 (47.83) | 0.70 (0.40-1.19) | 10 (58.82) | | 0.97 (0.49-1.86) |
| Poultry (meat) | 96 (41.03) | 9 (39.13) | 0.92 (0.52-1.57) | 5 (23.41) | | 0.65 (0.31-1.32) |
| Poultry (companionship) | 8 (3.42) | 1 (4.35) | 0.62 (0.45-4.40) | 2 (11.76) | | 3.42 (1.24-7.65) |
| Porcine (Fattening) | 99 (42.31) | 9 (39.13) | 0.85 (0.48-1.46) | 8 (47.06) | | 1.27 (0.63-2.44) |
| Porcine (piglet) | 52 (22.22) | 7 (30.43) | 1.48 (0.12-2.56) | 5 (29.41) | | 1.65 (0.82-3.19) |
| Porcine (companionship) | 3 (1.28) | 0 (0.00) | NS | 1 (5.88) | | NS |
| Turkeys (eggs) | 40 (17.09) | 6 (26.09) | 1.53 (0.80-2.75) | 2 (11.76) | | 0.54 (0.17-1.69) |
| Turkeys (meat) | 47 (20.09) | 5 (21.74) | 1.08 (0.54-2.04) | 3 (17.65) | | 0.89 (0.33-2.27) |
| Turkeys (companionship) | 2 (0.85) | 0 (0.00) | NS | 1 (5.88) | | 5.83 (1.46-2.15) |
| Packhorses | 47 (20.09) | 5 (21.74) | 1.07 (0.55-2.00) | 5 (29.41) | | 1.72 (0.83-3.38) |
| Horses (cowboy) | 27 (11.54) | 2 (8.70) | 0.77 (0.29-1.89) | 1 (5.88) | | 0.49 (0.12-1.89) |
| Horses (companionship) | 7 (2.99) | 0 (0.00) | NS | 1 (5.88) | | 2.13 (0.53-6.62) |
| Pack donkeys | 73 (31.20) | 6 (26.09) | 0.74 (0.39-1.36) | 9 (52.94) | | 2.39 (3.12-4.37) |
| Pack mules | 31 (13.25) | 0 (0.00) | NS | 2 (11.76) | | 0.90 (0.32-2.37) |
| **Synanthropic mammals** | | | | | | |
| Rats | 178 (76.07) | 21 (91.30) | 2.78 (1.13-6.29) | 13 (76.47) | | 0.94 (0.43-1.97) |
| **Practices common among families in study area** | | | | | | |
| Forest fragmentation and deforestation | 144 (61.54) | 14 (60.87) | 0.98 (0.56-1.67) | 12 (70.59) | | 1.42 (0.70-2.79) |
| Use of any rodent’s elimination measure | 131 (55.98) | 10 (43.48) | 0.70 (0.40-1.19) | 11 (64.71) | | 1.50 (0.73-2.95) |
